# Age differential analysis of COVID-19 second wave in Europe reveals highest incidence among young adults

**DOI:** 10.1101/2020.11.11.20230177

**Authors:** Alberto Aleta, Yamir Moreno

## Abstract

Most of the western nations have been unable to suppress the COVID-19 and are currently experiencing second or third surges of the pandemic. Here, we analyze data of incidence by age groups in 25 European countries, revealing that the highest incidence of the current second wave is observed for the group comprising young adults (aged 18-29 years old) in all but 3 of the countries analyzed. We discuss the public health implications of our findings.

## Body

Europe is experiencing a second wave of the COVID-19 pandemic, with incidence levels rising across all European countries. Authorities have already begun to enforce non-pharmaceutical interventions (NPIs) to avoid the collapse of the healthcare system, which will likely induce further drops in the estimated 2020 Gross Domestic Product due to additional slowdowns of the economy in many sectors. Whether we are able to tailor our response to this second wave in such a way that NPIs minimize social impact and economical losses is key for the future of Europe and its rapid recovery after the pandemic is brought under full control. This fundamentally depends on understanding better how the current surge of COVID-19 is unfolding and affecting the different strata of the population, which will eventually make it possible to adapt public policy responses and implement targeted measures on the fly.

To understand the evolution of the ongoing wave, one should trace it back to its origin in the summer, when the incidence started to grow again after the effects of the strict restrictions that were imposed in the Spring faded out, which resulted in an increased number of local outbreaks and community transmission of COVID-19 in most of Europe. For instance, it has been reported that a variant of SARS-CoV-2 emerged in early summer 2020 in Spain and spread to multiple European countries since then [1]. The source of the outbreak that Spain experienced in the summer can be related to outbreaks that started among agricultural workers in the north-east of the country - particularly in Aragon and Catalonia. These outbreaks then moved to the local population and replicated through the rest of the country. An analysis of the incidence by age group in one of these regions - Aragon, see Figure 1 - shows that in early summer the disease spread mainly within the 15-24 and 25-34 age groups. However, coinciding with the end of the summer and the start of the academic year, the disease spread mostly in the 15-24 age group. To elucidate if this pattern is characteristic of this region or if it is more general, we have collected data on the COVID19 incidence in 25 European countries aggregated by age groups during the period from 1 September to 27 October.

**Figure 1:**
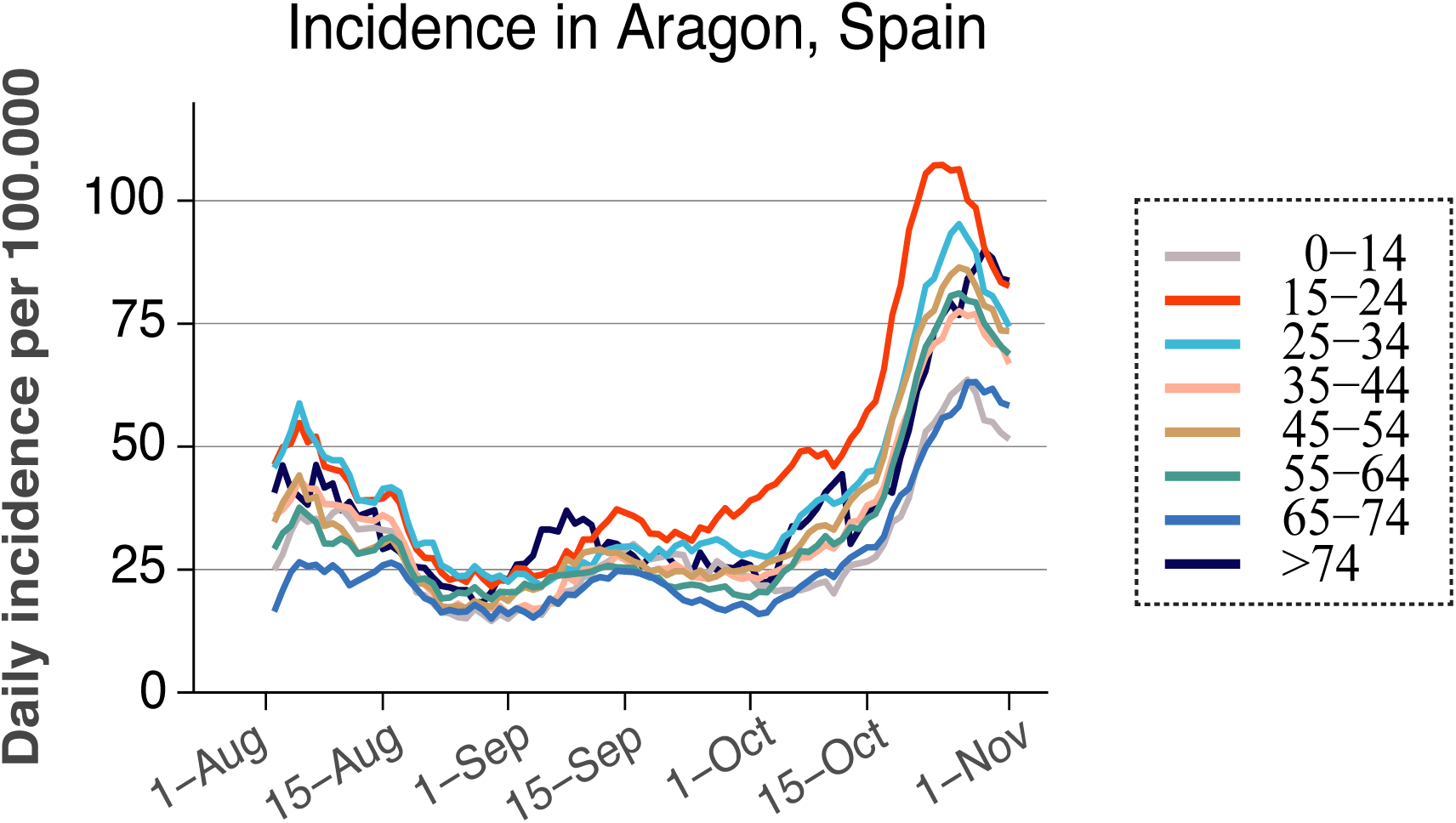
Daily incidence per age group in the region of Aragon, Spain.

Figure 2 shows the fraction of new cases in a given age bracket relative to the population of that age group. The horizontal line represents the situation in which the disease propagates homogeneously through the different age strata. Several features of the data are worth highlighting. First, in all countries but three (Czechia, Romania, and Slovenia), the maximum ratio of accumulated incidence in the period analyzed corresponds to the age bracket of young adults -mainly between 18-29 years old. Secondly, the incidence curves are not markedly peaked for the elderly as it happened in March-April during the first wave [2,3]. Third, the overall ratio of infected to population size of a given age group is below one for children in all countries. These very robust and regular patterns of the incidence of the current second wave are a remarkable observation given that countries in Europe responded distinctly and at different times during the early stages of the pandemic. The fact that the age-differential incidence is common to most countries in Europe points to common causes and similar transmission routes. Noticeably, a similar pattern has also been observed in the United States [4].

**Figure 2:**
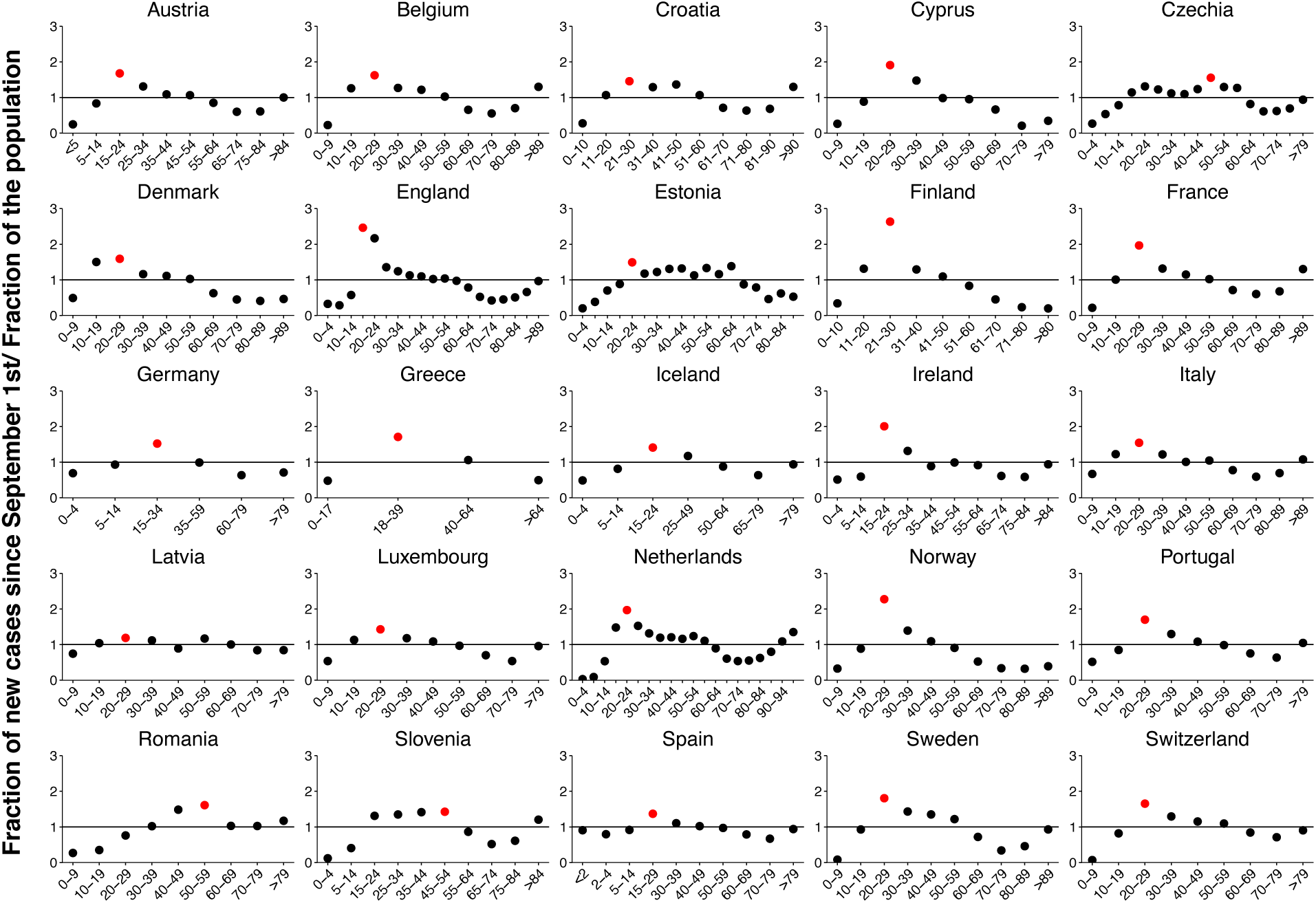
total number of new cases since 1 September to 27 October in each age bracket, divided by the size of the group. In red, the age bracket with the largest deviation from 1.

Several hypotheses could be advanced as most likely explanations for the striking regularity in age differential patterns across European countries. The higher incidence in young adults could result from interventions such as reopening of universities and other educational centers across Europe [5]. Nonetheless, it could also be rooted in behavioral factors associated with this age group [6], rather than from demographic or cultural causes specific to each European country or to differences in public interventions. These behavioral determinants could be the perception of low risk in this group and youngers’ social mixing and lifestyle -this is arguably the age group with the largest mobility and contact heterogeneity. The latter hypothesis is also consistent with the observation that the high incidence in younger people does not propagate to other age groups.

From a public health perspective, these data could prove fundamental to understand which factors impact the evolution of the current surge of COVID-19 and to adapt our response to the present unfolding of the COVID-19 pandemic. First, more efforts should be devoted to communicating with young adults to induce a change that reduces the incidence among them. Reporting data of incidence by age group to the general public could help to raise science-rooted awareness among the targeted age group. Secondly and important for the current debate regarding the role of schools, children seem to be infected less often than what would be expected if the transmission of SARS-CoV-2 would be homogeneously distributed across age groups. This points to the success of policies aimed at protecting this group [7,8]. We note that it is also possible that the observed pattern for the children is either because they could be less susceptible to be infected or that they might be less likely to be tested as they are mostly asymptomatic. Data however seems to indicate that the main reason is the former rather than the latter. Admittedly, if a large fraction of children were transmitting the disease, a surge in the age groups of their parents -those in close contact with them-should be expected, which is not a feature of the data in most countries. Likewise, the results suggest that schools for under 16-18 years old could remain unsheltered given that the benefits of keeping them open seem to overcome the social cost of a closure [9,10].

These results could help understand what are the main drivers of the second wave and to better design and adapt public health interventions during this stage of the pandemic. Furthermore, the previous findings highlight the need for more research in relation to tracing and identification of transmission chains, which will disambiguate what are the sources of contagion by age-strata. We also urge authorities to make available as much epidemiological data by age, sex, and other traits as possible to enable analyses like the one discussed here.

## Data Availability

Data used in this work is available from the authors.

